# Digital Phenotyping of Rest-Activity Rhythms and Biological Aging from Longitudinal Monitoring with Commercial Wearable Devices in *All of Us*

**DOI:** 10.1101/2025.09.26.25336772

**Authors:** Jinjoo Shim, J.P. Onnela

## Abstract

Poor circadian health is increasingly recognized as a determinant of aging and chronic diseases, yet longitudinal evidence in free-living populations remains limited. Most prior studies have been restricted to cross-sectional designs or short 7-day monitoring, precluding insight into long-term aging dynamics. To address this gap, we analyzed multi-year consumer wearable data linked with electronic health records from the *All of Us* Research Program to evaluate circadian rest-activity rhythms as longitudinal predictors of biological aging. Among 2,222 participants (median age 60.6 years, 68.5% female) contributing 8,447 person-years of Fitbit activity data with annual biological age estimates (PhenoAge), we performed high-dimensional digital phenotyping integrating functional data analysis with conventional rhythm metrics. Higher rhythm intensity reduced the odds of accelerated aging by 26-46%, greater regularity lowered the odds by 9-13%, whereas delayed acrophase increased the odds by 22%. Sex-stratified analyses revealed universal protection from rhythm intensity in both sexes, but stronger timing- and regularity-related vulnerabilities to accelerated aging in females (12-18% higher odds). In contrast, males exhibited a biphasic instability phenotype, characterized by early-morning surges and late-evening rebounds, uniquely linked to accelerated aging. This study provides the first large-scale longitudinal evidence establishing circadian rest-activity rhythms derived from consumer wearables as digital biomarkers of aging trajectories. With the growing scalability and ubiquity of consumer devices, our findings pave the way toward scalable aging risk assessment, targeted interventions, and advancing digital precision medicine to promote healthy longevity at the population level.

## INTRODUCTION

Circadian rhythms are fundamental biological processes that regulate sleep-wake cycles, metabolism, cognition, and immune function^1–4^. They align internal physiology with the external environment by shaping the amplitude and timing of daily functions to optimize health and survival. With aging, circadian organization deteriorates, manifested as dampened amplitude, increased fragmentation, and phase shifts^5,6^. While these changes are largely driven by intrinsic impairments in rhythmicity that perturbs cellular functions and the hormone regulation, circadian disruption may also occur by extrinsic stressors such as shift work, social jetlag, nighttime light exposure, and irregular activity schedules^7,8^. Thus, both intrinsic aging processes and extrinsic challenges progressively undermine circadian integrity, with downstream consequences for health and longevity.

Accumulating evidence links circadian disruption to a broad spectrum of adverse outcomes, including metabolic dysfunction, cardiovascular disease, neurodegeneration, mental illness, frailty, and elevated mortality risk^2,9–14^. More recently, circadian disruption has been recognized as a correlate of accelerated biological aging, a measure of physiological and functional state that more accurately predicts morbidity and mortality than chronological age^15–18^. Cross-sectional studies consistently demonstrate that attenuated amplitude, delayed acrophase, and irregular or fragmented activity rhythms are associated with reduced lifespan and healthspan. Yet, most evidence comes from single, short-duration activity recordings, and the longitudinal influence of circadian rhythmicity on biological aging trajectories remains unresolved.

This limitation reflects methodological constraints. Conventional circadian assessments such as melatonin profiling require repeated invasive, labor-intensive sampling and are impractical for large-scale studies^19^. As an alternative, research-grade actigraphy has been implemented in large cohort studies to characterize rest-activity rhythms, sleep-wake patterns, and physical activity in a non-invasive and continuous manner^20–22^. However, a majority of studies using wearable actigraphy rely on 7-day recordings, providing only a limited snapshot of activity patterns. Such short assessments are also highly susceptible to transient influences such as acute illness, travel, environmental factors, or seasonal variation^23^. These constraints hinder the identification of persistent circadian rhythm features that could serve as robust and modifiable biomarkers of biological aging.

Recent advances in digital phenotyping and widespread adoption of consumer-grade digital devices now offer a way forward^24,25^. Wearables and smartphones allow passive, continuous, and long-term monitoring of rest-activity rhythms, sleep, and activity in free-living settings, enabling scalable longitudinal characterization^26,27^. This creates a unique opportunity to integrate repeated measures of biological age assessments with digital phenotyping of real-world rest-activity rhythm dynamics, thereby capturing persistent circadian features that short-term actigraphy is limited to detect^28,29^.

To this end, we leveraged the *All of Us* Research Program, a large nationwide prospective cohort in the United States integrating multi-year, multi-modal data to examine the relationship between longitudinal wearable-based activity rhythms and biological age acceleration. Using Fitbit activity data, we extracted circadian rest-activity rhythm metrics over time and applied data-driven functional principal component analysis to characterize latent 24-hour rhythm profiles. By linking these longitudinal activity rhythm profiles with repeated measures of biological age, quantified by phenotypic age (PhenoAge)^30^, we provide the first large-scale evidence that persistent circadian rhythmicity functions as a digital biomarker of aging risk. Our findings not only establish rest-activity rhythm as a scalable and accessible biomarker of aging but also lay the groundwork for precision-prevention strategies targeting circadian health to promote healthy longevity.

## RESULTS

### Baseline characteristics

**Table 1** describes the baseline characteristics of the study cohort (N=2,222), stratified by biological aging status. Of the participants, 1,021 (45.9%) were classified as fast agers and 1,201 (54.1%) as slow agers at baseline. Chronological age did not differ significantly between the groups (61.09 years, SD 13.82 vs. 60.23 years, SD 14.17; two-sample *t-*test of Δ≠0: *p*=0.147), whereas biological age was significantly higher in fast agers (66.43 years, SD 14.57) compared with slow agers (54.34 years, SD 14.48, two-sample *t-*test of Δ≠0: *p*<0.001).

**Table 1.** Baseline characteristics of the study population by biological aging status.

| Mean (SD) or n (%) | All<br>(n=2222) | Fast Aging<br>(n=1021) | Slow Aging<br>(n=1201) | p-value |
| --- | --- | --- | --- | --- |
| Age, years | 60.62 (14.01) | 61.09 (13.82) | 60.23 (14.17) | 0.147 |
| PhenoAge, years | 59.90 (15.72) | 66.43 (14.57) | 54.34 (14.48) | <0.001 |
| Sex |  |  |  | <0.001 |
| Female | 1522 (68.5) | 636 (62.3) | 886 (73.8) |  |
| Male | 693 (31.2) | 380 (37.2) | 313 (26.1) |  |
| Race |  |  |  | 0.008 |
| White | 1839 (82.8) | 838 (82.1) | 1001 (83.3) |  |
| Black | 134 (6.0) | 78 (7.6) | 56 (4.7) |  |
| Other | 249 (11.2) | 105 (10.3) | 144 (12.0) |  |
| BMI, kg/m<sup>2</sup> | 29.84 (6.98) | 31.76 (7.48) | 28.21 (6.07) | <0.001 |
| Education |  |  |  | 0.187 |
| College or above | 2084 (93.8) | 958 (93.8) | 1126 (93.8) |  |
| 12 years or GED | 119 (5.4) | 50 (4.9) | 69 (5.7) |  |
| Income |  |  |  | <0.001 |
| High | 1015 (45.7) | 428 (41.9) | 587 (48.9) |  |
| Medium | 669 (30.1) | 303 (29.7) | 366 (30.5) |  |
| Low | 395 (17.8) | 224 (21.9) | 171 (14.2) |  |
| Missing | 143 (6.4) | 66 (6.5) | 77 (6.4) |  |
| Employment |  |  |  | 0.011 |
| Employed | 1260 (56.7) | 564 (55.2) | 696 (58.0) |  |
| Retired | 732 (32.9) | 345 (33.8) | 387 (32.2) |  |
| Unemployed | 159 (7.2) | 88 (8.6) | 71 (5.9) |  |
| Other | 71 (3.2) | 24 (2.4) | 47 (3.9) |  |
| Smoking |  |  |  | 0.048 |
| Yes | 788 (35.5) | 394 (38.6) | 394 (32.8) |  |
| No | 1434 (64.5) | 627 (61.4) | 807 (67.2) |  |
| Drinking |  |  |  | 0.36 |
| Yes | 2168 (97.6) | 1000 (97.9) | 1168 (97.3) |  |
| No | 54 (2.4) | 21 (2.1) | 33 (2.7) |  |
| Comorbidities |  |  |  |  |
| Cancer | 207 (9.3) | 92 (9.0) | 115 (9.6) | 0.702 |
| CVD | 247 (11.1) | 107 (10.5) | 140 (11.7) | 0.417 |
| Type 2 diabetes | 212 (9.5) | 97 (9.5) | 115 (9.5) | 1.000 |
| Daily steps | 7715.83<br>(3429.04) | 7215.22<br>(3374.21) | 8141.42<br>(3419.17) | <0.001 |
| Sleep duration, hours | 6.39 (1.67) | 6.32 (1.67) | 6.46 (1.66) | 0.048 |

Values are mean (SD) for continuous variables and n (%) for categorical variables. *p*-values are based on *t*-tests or chi-square tests, as appropriate.

Compared to slow agers, the fast agers group had a greater proportion of males (37.2% vs. 26.1%, *p*<0.001) and a higher mean body mass index (BMI: 31.76 kg/m² vs. 28.21 kg/m²; *p*<0.001). Socioeconomic indicators showed consistent differences as low income was more prevalent among fast agers (21.9% vs. 14.2%; *p*<0.001), as was unemployment (8.6% vs. 5.9%; *p*=0.011). Fast agers also had a higher prevalence of lifetime smoking (38.6% vs. 32.8%; *p*=0.005). In line with behavioral differences, fast agers showed shorter average sleep duration (6.32 hours vs. 6.46 hours; *p*=0.048) and lower wearable-derived daily step counts (7,215 steps vs. 8,141 steps; *p*<0.001). Education, current alcohol consumption, and comorbidities of cancer, cardiovascular disease (CVD), and diabetes did not differ significantly between the groups.

PhenoAge was strongly correlated with chronological age (Pearson correlation coefficient, *r*=0.87, *p*<0.001) in the full sample, with consistent associations observed in males (*r*=0.85, *p*<0.001) and females (*r*=0.88, *p*<0.001) (**Supplemental Fig. 1**). The distribution of biological age acceleration (ΔAge) approximated normality and was centered near zero. Sex-specific distributions of ΔAge were comparable in terms of central tendency and dispersion.

Baseline rest-activity profiles by biological aging status (**Supplemental Fig. 2**) further highlighted rhythmicity distinctions. Across age strata, fast agers exhibited consistently lower peak activity levels and attenuated daytime intensity compared to slow agers. They also showed higher nocturnal activity between 00:00-06:00, whereas slow agers maintained a more consolidated rest period, consistent with a more robust circadian rhythm.

### Age-related trends in circadian rest-activity rhythms (CRAR) and functional components

**Fig. 1** illustrates age-related trends in Circadian Rest-Activity Rhythm (CRAR) metrics. Across age ranges, slow agers steadily exhibited higher MESOR (a rhythm-adjusted mean), cosinor amplitude, RA (relative amplitude), M10 (mean activity during the 10 most active hours), and IS (interdaily stability; day-to-day rhythm regularity) compared with fast agers.

**Fig. 1.**
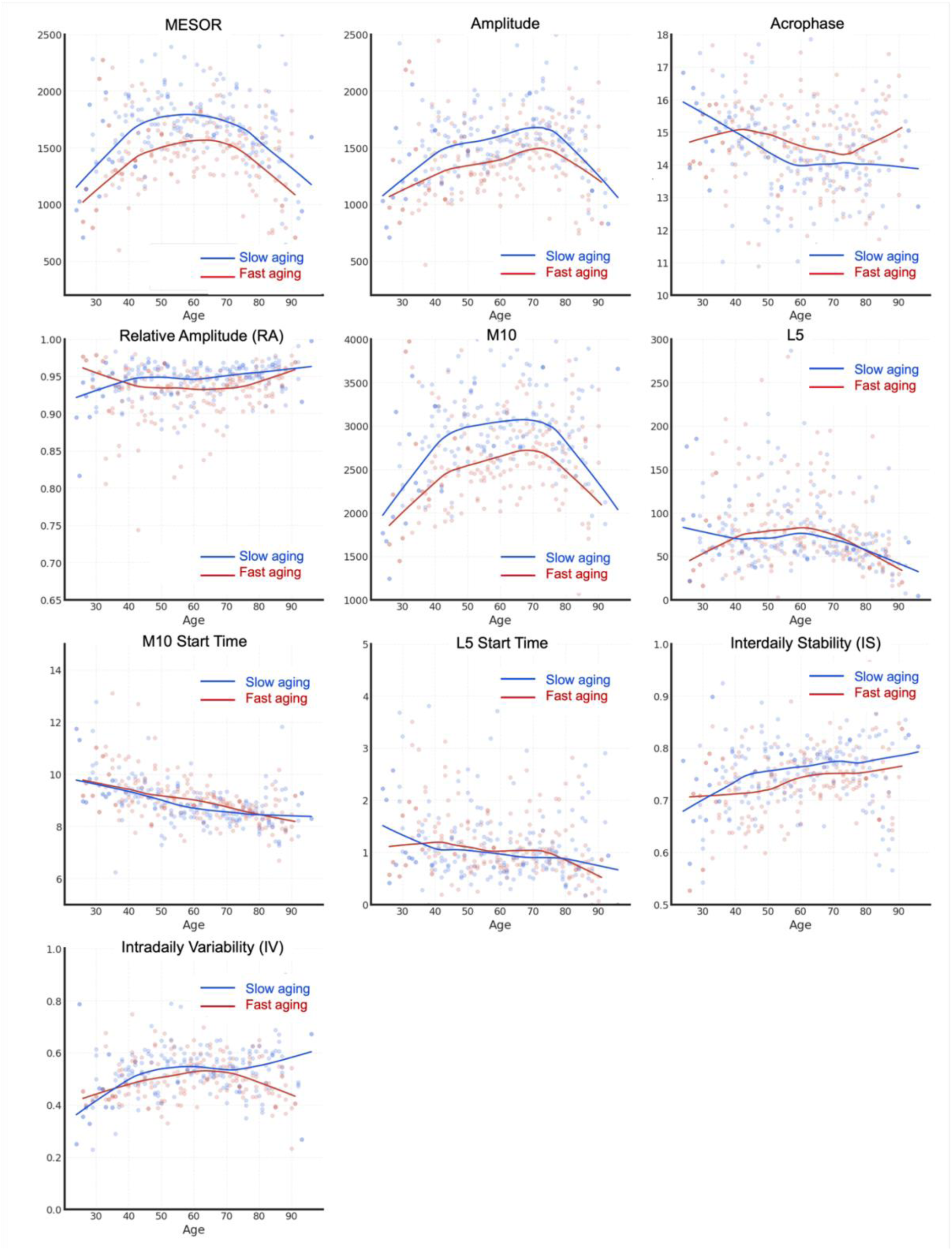
Age-associated trends in circadian rest-activity rhythm (CRAR) metrics by biological age acceleration. Points are color-coded by biological aging status at baseline (blue=slow aging, red=fast aging). Locally weighted scatterplot smoothing lines are overlaid to visualize smoothed trajectories by aging group.

Differences in acrophase emerged from midlife (∼40 years) onward, with fast agers showing delayed acrophase. Sex-stratified analyses confirmed the robustness of these trends (**Supplementary** Fig. 3). In both sexes, fast aging was linked to lower MESOR, amplitude, M10, and IS, but the magnitude of the difference varied. In female, differences remained stable with age, whereas in male they attenuated after 60. For L5 (mean activity during the least active hours), differences were more pronounced in male.

Functional principal component analysis (fPCA) captured latent nonlinear variation in 24-h profiles (**Fig. 2**). The first four fPCs accounted for 88% of total variance. fPC1 (accounting for 51% of the variance) quantified overall daytime activity amplitude, with higher scores indicating greater activity levels. fPC2 (18% of the variance) represented the timing of activity onset, where lower scores reflected delayed transitions from rest to activity. fPC3 (13% of the variance) described the timing of peak activity, differentiating morning- from evening-dominant patterns. fPC4 (6% of the variance) distinguished biphasic from monophasic activity structures.

**Fig. 2.**
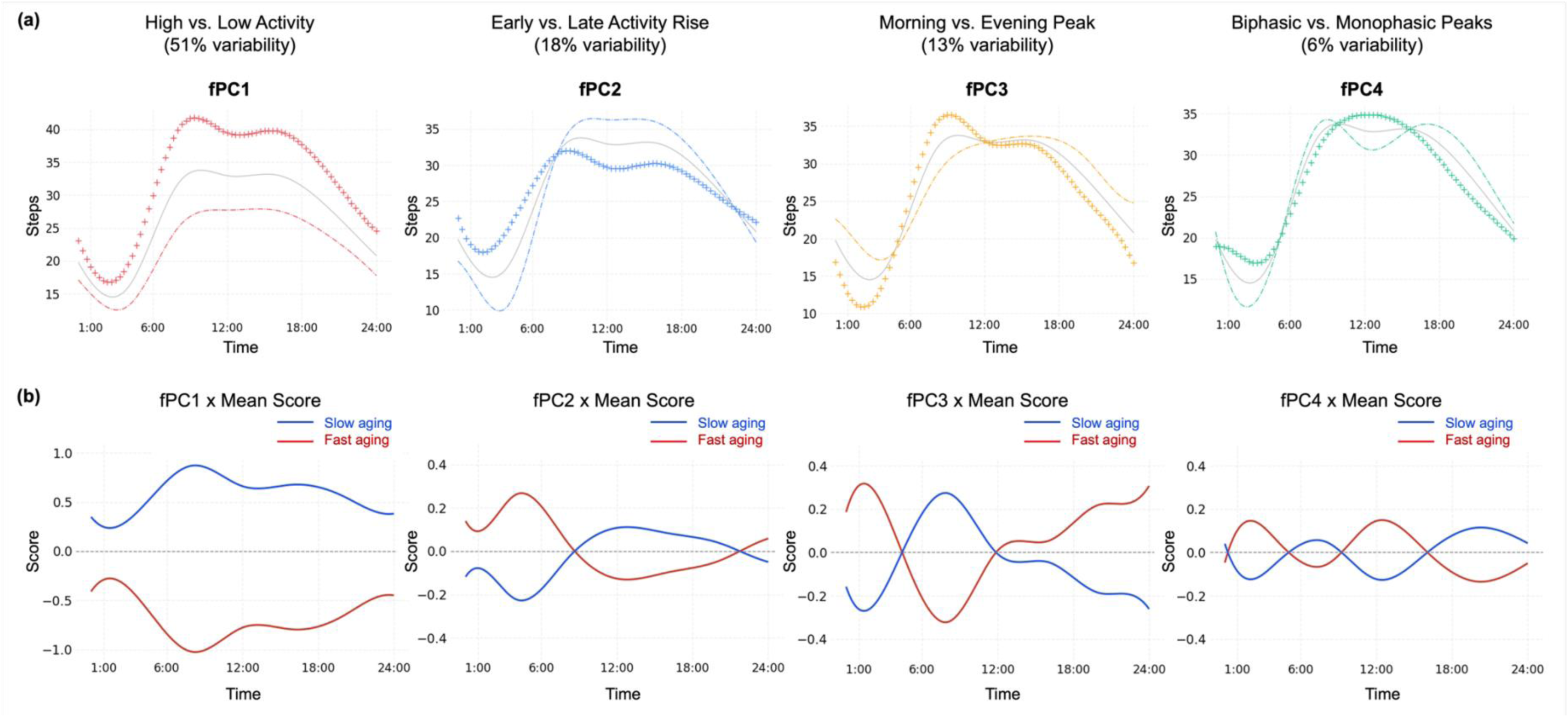
**Functional principal components (fPCs) of 24-hour activity profiles and their association with biological age acceleration**. **(a)** Mean 24-hour activity profiles are plotted separately by the sign of each fPC score, illustrating the predominant temporal patterns captured by each component. **(b)** Line plots depict the fPC mean score within each biological aging group at baseline (blue=slow aging, red=fast aging).

Group-level comparisons revealed distinct rhythm signatures of fast aging. Fast agers exhibited lower fPC1 scores reflecting reduced daytime activity. fPC2 trajectories indicated delayed activity rise with blunted midday recovery. fPC3 revealed late-evening activity peaks, which suggests circadian misalignment or shifted rhythms. fPC4 indicated biphasic activity with an early-morning and late-day peaks. Correlation analyses (**Supplementary** Fig. 4) confirmed fPC1 aligned with MESOR, amplitude, RA, and M10, underscoring its representation of rhythm intensity. fPC2 and fPC3 correlated with timing metrics such as acrophase, M10 start time, and L5 start time.

Sex-specific fPC profiles sharpened these patterns (**Supplementary** Fig. 5). Female showed more pronounced timing-related shifts (fPC2, fPC3), while male displayed distinct fPC4 separation, with fast-aging male exhibiting bimodal rhythms and increased nocturnal activity. Together, CRAR metrics and fPCs delineate a fast-aging phenotype characterized by reduced amplitude, delayed activity onset/peak, and rhythm fragmentation, with sex-specific nuances.

### Associations between functional and circadian rhythm features and accelerated biological aging at baseline and follow-up

We next assessed the associations of rhythmicity with accelerated biological aging at baseline and 5-year follow-up (**Table 2**). Across both time points, higher rhythm intensity (fPC1, MESOR, amplitude, RA, M10) and greater rhythm regularity (IS) reduced the odds of accelerated aging, whereas delayed timing metrics (acrophase, M10 start time, L5 start time) increased the odds. For example, each quartile increase in fPC1 corresponded to 19.0% lower odds of accelerated aging at baseline (OR=0.810; 95% CI: 0.745-0.878) and a 19.4% reduction at follow-up (OR=0.806; 95% CI: 0.697-0.933). Comparable protective effects were observed for MESOR (baseline OR=0.778; follow-up OR=0.787), amplitude (baseline OR=0.781; follow-up OR=0.831), RA (baseline OR=0.829; follow-up OR=0.889), and M10 (baseline OR=0.762; follow-up OR=0.779).

**Table 2.** Associations between functional and circadian rhythm features and accelerated biological aging at baseline and follow-up.

| **Features** | **Baseline OR (95% CI)** | **p-value** | **Follow-up OR (95% CI)** | **p-value** |
| --- | --- | --- | --- | --- |
| fPC1 | **0.810 (0.745, 0.878)** | **<0.001** | **0.806 (0.697, 0.933)** | **0.004** |
| fPC2 | 1.044 (0.965, 1.130) | 0.287 | 0.974 (0.853, 1.169) | 0.699 |
| fPC3 | 0.956 (0.884, 1.035) | 0.265 | 0.909 (0.799, 1.036) | 0.152 |
| fPC4 | 1.003 (0.927, 1.086) | 0.934 | 0.978 (0.857, 1.108) | 0.694 |
| MESOR | **0.778 (0.717, 0.845)** | **<0.001** | **0.787 (0.680, 0.910)** | **0.001** |
| Amplitude | **0.781 (0.718, 0.847)** | **<0.001** | **0.831 (0.721, 0.958)** | **0.011** |
| Acrophase | **1.109 (1.024, 1.200)** | **0.011** | 1.014 (0.887, 1.161) | 0.835 |
| Relative amplitude (RA) | **0.829 (0.764, 0.899)** | **<0.001** | 0.889 (0.776, 1.019) | 0.093 |
| M10 | **0.762 (0.703, 0.829)** | **<0.001** | **0.779 (0.674, 0.901)** | **0.001** |
| L5 | 1.047 (0.966, 1.134) | 0.266 | 1.018 (0.891, 1.163) | 0.795 |
| M10 start time | 1.066 (0.983, 1.155) | 0.124 | **1.149 (1.001, 1.318)** | **0.048** |
| L5 start time | **1.106 (1.021, 1.198)** | **0.014** | 1.122 (0.982, 1.280) | 0.09 |
| Interdaily stability (IS) | **0.889 (0.820, 0.964)** | **0.004** | 0.879 (0.768, 1.006) | 0.061 |
| Intraday variability (IV) | 0.938 (0.866, 1.016) | 0.116 | 0.910 (0.795, 1.042) | 0.172 |

Estimates are derived from generalized linear models adjusted for age, sex, race/ethnicity, education, income, employment, lifetime smoking, current alcohol consumption, and comorbidities (cancer, CVD, type 2 diabetes). Significant associations ( $p < 0.05$ ) are shown in bold.

Higher rhythm regularity (IS) demonstrated reduced odds of accelerated aging at baseline (OR=0.889; 95% CI: 0.820-0.964) and follow-up (OR=0.879; 95% CI: 0.768-1.006).

In contrast, delayed acrophase (peak activity timing) and late L5 start time (the least active 5-hour onset timing) increased baseline odds by 10.9% (OR=1.109; 95% CI: 1.024-1.200) and 10.6% (OR=1.106; 95% CI: 1.021-1.198), respectively, and delayed M10 start time (the most active 10-hour onset timing) raised follow-up odds by 14.9% (OR=1.149; 95% CI: 1.001-1.318).

To determine whether rhythm-aging associations are stable over time, we examined group-level trajectories of circadian metrics at years 1, 3, and 5 (**Fig. 3**) and estimated adjusted odds ratios for accelerated aging across the observed metric ranges (**Fig. 4**). The direction and magnitude of associations remained largely consistent, with the strongest effects observed at year 1 and modest attenuation thereafter, reinforcing the temporal stability of these relationships. Sex-stratified analyses (**Supplementary Table 1**) showed overall consistency, though timing effects were more pronounced in female, while rhythm intensity remained protective across both sexes.

**Fig. 3.**
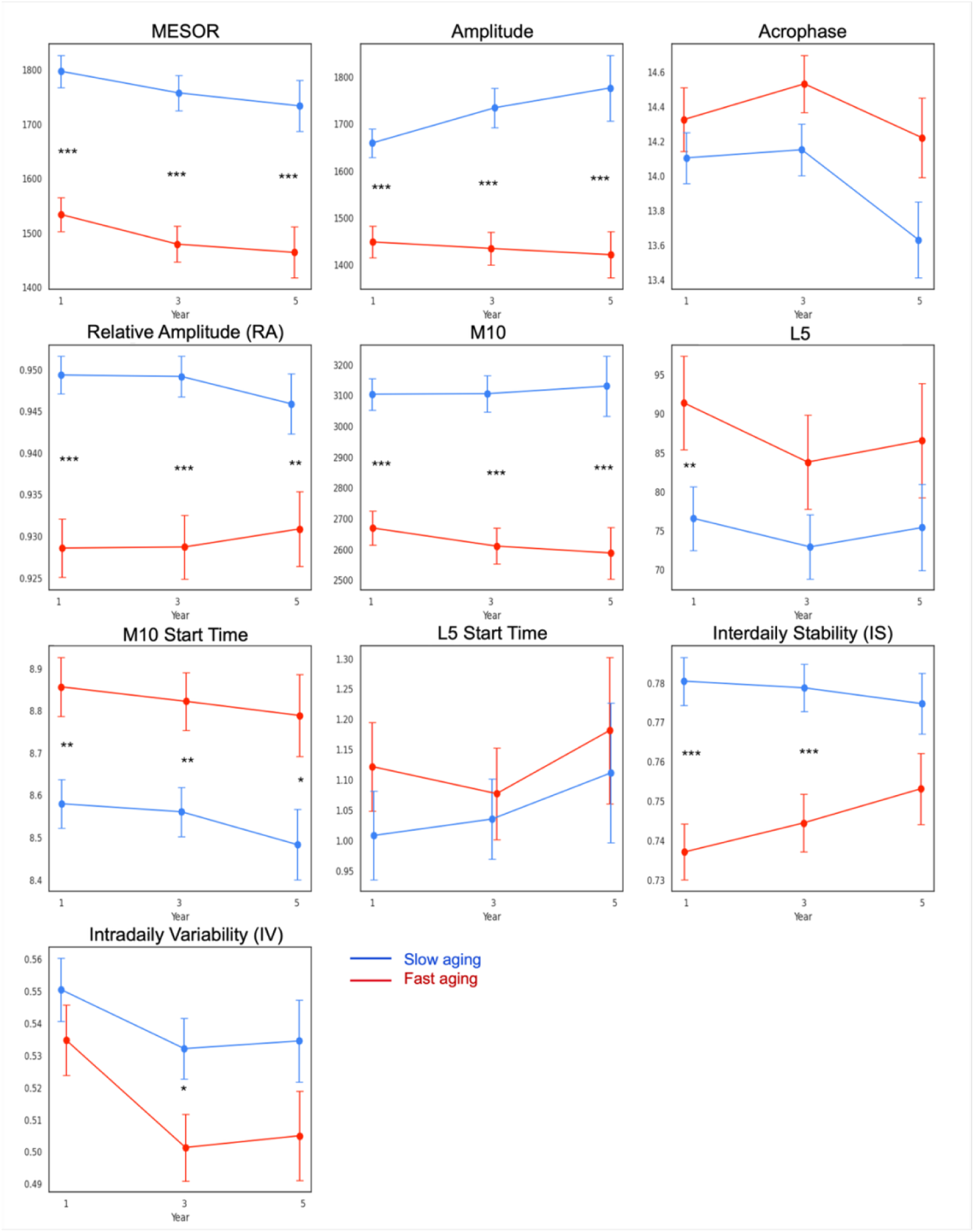
Trajectories of circadian rhythm metrics over 5 years stratified by biological age acceleration. Mean values (± standard error) are shown for fast agers (red) and slow agers (blue), classified according to biological age acceleration at each follow-up. Asterisks denote the statistical significance of between-group differences at each time point (\**p*<0.05, \*\**p*<0.01, \*\*\**p*<0.001).

**Fig. 4.**
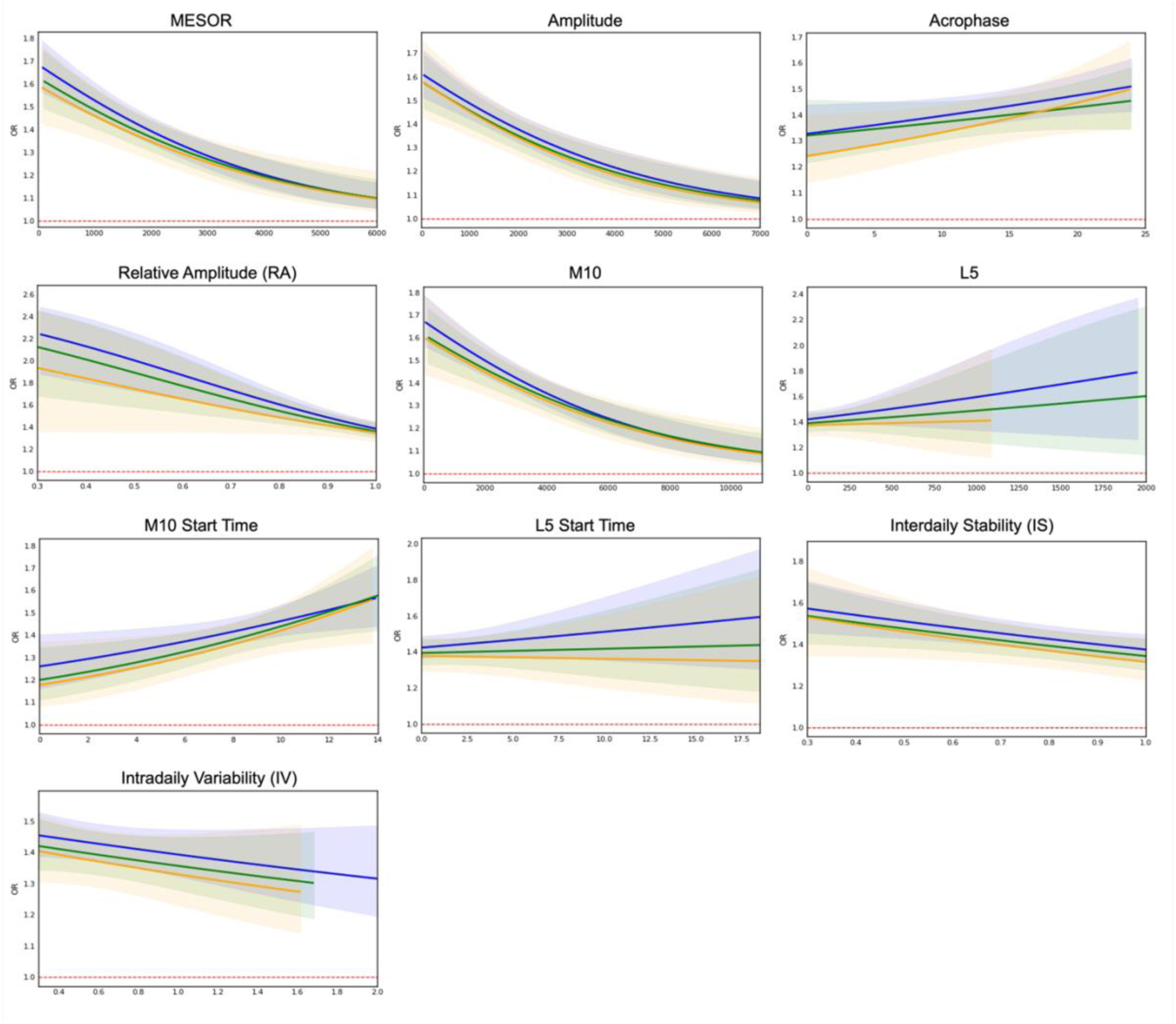
**Associations between functional and circadian rhythm features and accelerated biological aging stratified by years**. Each panel displays the adjusted odds ratio (OR) for fast aging across the observed range of a given rhythm metric stratified by study year. Blue, green, and orange lines correspond to data from Year 1, Year 3, and Year 5, respectively. Shaded regions represent 95% confidence intervals. All models are adjusted for age, sex, race/ethnicity, education, income, employment, lifetime smoking, current alcohol consumption, and comorbidities (cancer, CVD, type 2 diabetes). The red dashed horizontal line at OR=1 denotes the null effect reference. ORs are estimated over the observed data range while covariates are held at reference (mode or mean) values.

### Longitudinal associations of functional and circadian rhythm metrics with biological aging

To move beyond cross-sectional contrasts, we evaluated whether changes in rhythm predicted changes in biological aging over time (**Table 3**). Linear mixed-effects models demonstrated robust longitudinal associations, accounting for within-subject repeated measurements. Higher activity intensity and rhythm robustness reduced odds of accelerated aging across the study period. A one-quartile increase in fPC1 corresponded to a 36.9% reduction in the odds (OR=0.631; 95% CI: 0.526-0.757; *p*<0.001). Similarly, increases in MESOR, amplitude, RA, and M10 were linked to 42.4% (OR=0.576; 95% CI: 0.475-0.699), 40.2% (OR=0.598, 95% CI: 0.494-0.723), 26.5% (OR=0.735, 95% CI: 0.610-0.884), and 46.3% (OR=0.537, 95% CI: 0.442-0.652) reductions, respectively (all *p*<0.001). Increased rhythm regularity showed 9% reduced odds (OR=0.910, 95% CI: 0.756-1.095).

**Table 3.** Longitudinal associations of functional and circadian rhythm metrics with biological aging over time.

| Features | OR (95% CI) | p-value |
| --- | --- | --- |
| fPC1 | **0.631 (0.526, 0.757)** | **<0.001** |
| fPC2 | 0.983 (0.825, 1.172) | 0.851 |
| fPC3 | 0.955 (0.801, 1.140) | 0.611 |
| fPC4 | 1.022 (0.857, 1.219) | 0.807 |
| MESOR | **0.576 (0.475, 0.699)** | **<0.001** |
| Amplitude | **0.598 (0.494, 0.723)** | **<0.001** |
| Acrophase | **1.221 (1.015, 1.468)** | **0.034** |
| Relative amplitude (RA) | **0.735 (0.610, 0.884)** | **0.001** |
| M10 | **0.537 (0.442, 0.652)** | **<0.001** |
| L5 | 0.974 (0.974, 1.175) | 0.788 |
| M10 start | 1.060 (0.878, 1.279) | 0.545 |
| L5 start | 1.082 (0.899, 1.302) | 0.405 |
| Interdaily stability (IS) | 0.910 (0.756, 1.095) | 0.319 |
| Intraday variability (IV) | 0.872 (0.723, 1.053) | 0.155 |

Odds ratios (ORs) and 95% confidence intervals (CIs) are derived from linear mixed-effects regression models. Models include a random intercept and adjust for age, sex, race/ethnicity, education, income, employment, lifetime smoking, current alcohol consumption, and comorbidities (cancer, CVD, type 2 diabetes). Significant associations ( $p<0.05$ ) are shown in bold.

In contrast, delayed acrophase increased odds by 22.1% (OR=1.221; 95% CI: 1.015-1.468; *p*=0.034), while later M10 and L5 start timings showed modest, non-significant increases by 6.0% (OR=1.060; 95% CI: 0.878-1.279) and 8.2% (OR=1.082; 95% CI: 0.899-1.302), respectively.

Within-person analyses supported these findings (**Fig. 5**). Participants with increases in intensity metrics (MESOR, amplitude, RA, M10) demonstrated negative ΔAge slopes, indicating decelerated aging, whereas delays in acrophase and M10 start time produced positive ΔAge slopes, indicating accelerated aging trajectories. Dispersion of slopes was narrower for intensity metrics, suggesting more uniform protective effects across individuals, while timing effects were more heterogeneous.

**Fig. 5.**
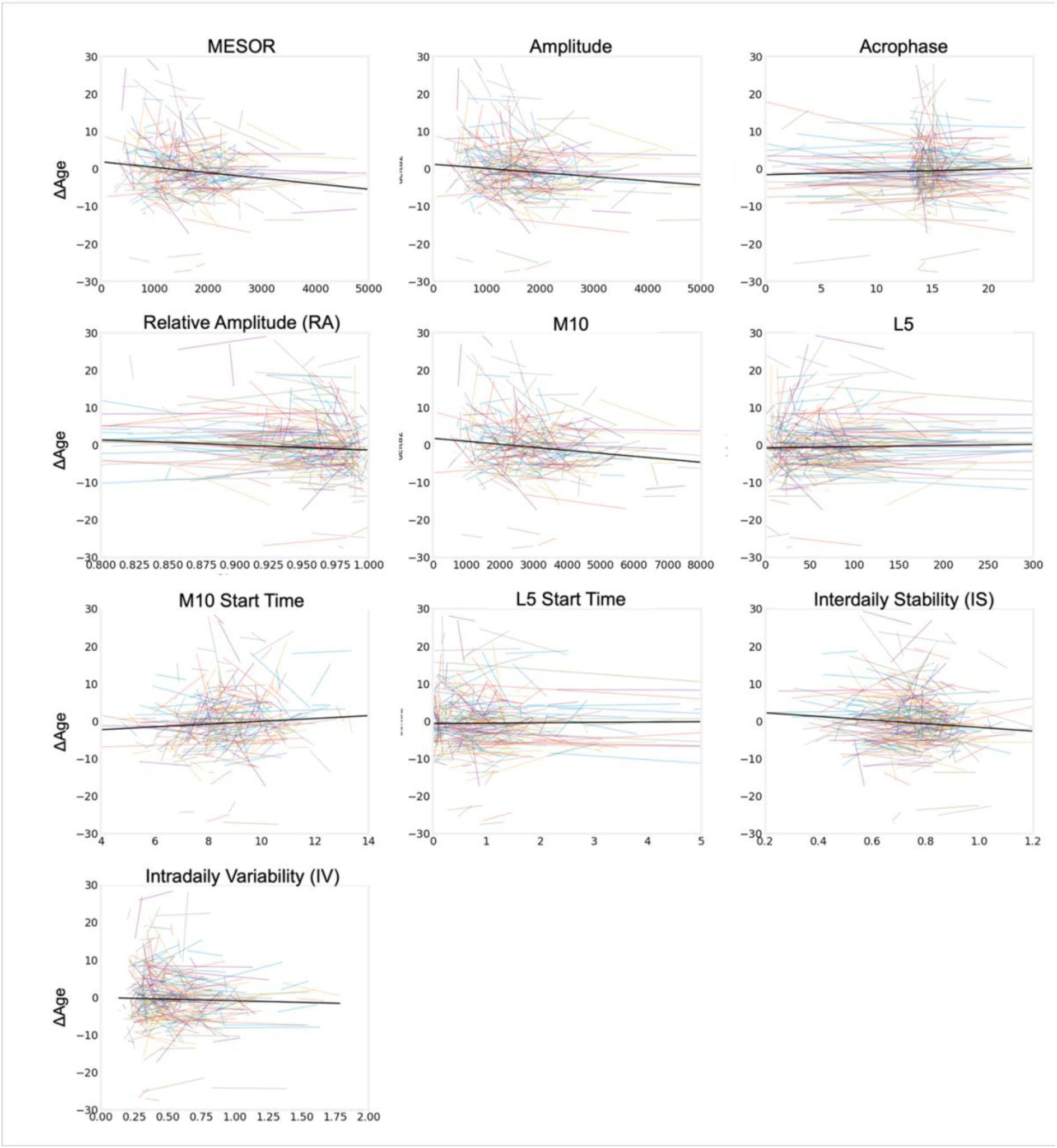
Within-individual changes in circadian rhythm metrics versus change in **biological aging.** Association between activity rhythm metrics and changes in biological age acceleration (ΔAge). Each panel shows the linear relationship between one wearable-derived feature (x-axis) and the within-person change in ΔAge (y-axis, in years). The colored lines represent individual-level regression slopes estimated for a random sample of 100 participants with ≥2 observations. The bold black line depicts the population-level trend estimated across the entire dataset.

Sex-stratified longitudinal analyses (**Supplementary Table 2**) confirmed these trends and highlighted sex-specific nuances. Intensity-related protection against accelerated aging was evident in both sexes. In male, higher fPC1, MESOR, amplitude, and M10 consistently reduced the odds of fast aging by 15-30%, whereas the effect sizes were larger in female, ranging from 22 to 40%. Increased rhythm regularity reduced the odds of accelerated aging by 15% in female, while showing no statistically significant effect in male. Similarly, delayed acrophase increased the odds of fast aging by 12% in male and 18% in female but was statistically significant only in female.

Together, the longitudinal analyses extend the cross-sectional findings by showing that within-individual changes in circadian rhythmicity dynamically shape the pace of biological aging, with intensity and robustness emerging as consistent predictors and timing and regularity exerting more sex-specific effects.

## DISCUSSION

In this large-scale, prospective cohort with multi-year wearable monitoring in free-living settings, we demonstrated that circadian rest-activity rhythms are robust determinants of biological aging trajectories. Rhythm intensity, regularity, and timing emerged as important predictors of accelerated aging at baseline and during follow-up. Longitudinal analyses indicated that changes in rest-activity rhythms prospectively shaped biological aging trajectories across five years. Specifically, higher rhythm intensity reduced the odds of accelerated aging by 26-46%, greater regularity decreased the odds by 9-13%, whereas delayed timing increased the odds by 22%. Rhythm intensity provided stable protection across both sexes, while reduced regularity and delayed timing were specific risk factors in females. Males exhibited a distinctive biphasic instability phenotype linked to accelerated aging. By leveraging repeated longitudinal assessments, these findings move beyond single time-point associations to provide large-scale longitudinal evidence establishing consumer wearable-derived circadian rhythms as digital biomarkers of biological aging. Given the increasing scalability and ubiquity of consumer wearables, this work shows the potential for continuous monitoring and near real-time interventions of aging to support digital precision medicine and promote healthy aging.

A distinctive contribution of this work lies in the integration of data-driven functional principal components analysis (fPCA) with conventional scalar circadian metrics, enabling high-dimensional digital phenotyping of rhythm intensity, regularity, and timing. fPCA revealed additional latent signatures characterizing accelerated aging, including decreased overall robustness (fPC1), delayed initiation (fPC2), peak misalignment (fPC3), and biphasic instability (fPC4). These functional dimensions demonstrated strong concordance with scalar measures while providing enhanced biological granularity. Notably, fPC1, reflecting rhythm robustness, was one of the most sensitive predictors of accelerated aging, underscoring the primacy of rhythm intensity as a protective factor. This demonstrates the value of multidimensional digital phenotyping for uncovering hidden vulnerabilities in circadian-aging biology.

Our results also extend prior evidence linking circadian disruption to health outcomes. Reduced amplitude has been consistently identified as a strong predictor of mortality, cardiovascular disease, cancer, and neurodegenerative disorders^15,31–34^, with more recent associations to preclinical conditions such as frailty^35^ and cognitive decline^36^.

Decrease in amplitude is also further tied to systemic inflammation (“inflammaging”), a hallmark of aging and a common pathway for multiple chronic diseases^3,37^, suggesting that weakening rest-activity rhythm may accelerate aging even at younger ages^38^. Our findings extend this by showing that, in longitudinal analyses, amplitude and rhythm intensity remain the strongest predictors of accelerated aging.

Timing-related disruption provides another pathway of vulnerability. Delayed acrophase or later activity onset has been associated with elevated risk of cardiovascular disease, obesity, cancer, and dementia^39–41^, though effect sizes and significance vary across studies. Chronic night shift work, an exemplar of sustained circadian misalignment, has been linked to higher brain age indices, greater prevalence of metabolic syndrome, and increased risk of atrial fibrillation and coronary heart disease^42–44^. These cumulative, persistent effects are often underestimated or missed in cross-sectional studies. Consistent with this, our findings demonstrated that delayed acrophase was prospectively associated with accelerated aging, implying the lasting impact of circadian timing disruption on long-term aging trajectories.

Furthermore, rhythm regularity adds complementary insights. While day-to-day regularity often increases with aging^45^, reduced stability and greater fragmentation have been associated with higher risk of neurodegenerative disease^46^. In our cohort, greater regularity provided protection against accelerated aging in females, whereas in males we identified a novel biphasic instability phenotype, characterized by early-morning surge and late-evening rebound, uniquely linked to accelerated aging.

Together, these results emphasize the importance of assessing rhythm amplitude, regularity, and timing as an integrated multidimensional framework rather than as isolated parameters. In this line of work, prior studies have shown that wearable-based circadian rhythm-based age outperforms physical activity-based age in predicting morbidity and mortality across cardiovascular, metabolic, and neurodegenerative diseases, reinforcing the value of comprehensive rhythm profiling^18^. Evidence from a cohort of 191 older adults further showed that rhythm intensity, regularity, and timing jointly associated with epigenetic age acceleration showing their interdependent effects^47^. By combining repeated measures of biological age acceleration and high-dimensional functional analyses, our study advances prior cross-sectional evidence and precisely establishes the longitudinal role of rest-activity rhythm patterns as robust digital biomarkers of aging trajectories.

Sex-stratified analyses further refined these insights by revealing differential vulnerabilities. Rhythm intensity was broadly protective in both sexes, but timing- and regularity-related associations were markedly stronger in females, where delayed acrophase and later rest onset increased the odds of accelerated aging by 12-18% and greater regularity reduced risk by 15%. In males, timing-related effects were weaker, yet fPCA identified the biphasic instability phenotype as a unique marker of accelerated aging.

Several mechanisms may explain these sex-specific pathways. First, aging is accompanied by intrinsic weakening of circadian amplitude, reduced output from the suprachiasmatic nucleus (SCN), and impaired peripheral clock coordination. Importantly, the SCN can regulate circadian phase and rhythm in a sex-specific manner through hormonal modulation, behavioral differences in rest-activity and sleep quality, and immunological dimorphisms^48–50^. For example, estrogen enhances pacemaker sensitivity and circadian amplitude, potentially accounting for the stronger phase-related effects observed in female. In addition, female generally exhibit more robust and stable circadian rhythms, whereas male more often display rhythm fragmentation and inconsistency^45^ and lower sleep efficiency^51^. Under real-world conditions, male also tend to have later chronotypes^52^, predisposing them to higher rates of social jetlag^52^ and associated circadian disruption.

Collectively, these results suggest that circadian aging trajectories diverge by sex, shaped by an interplay of intrinsic molecular regulation and behavioral-environmental exposures. It is also noteworthy that the relatively smaller number of males in our cohort likely reduced statistical power, attenuating associations in males.

The translational implications are considerable. Our dual-domain framework, highlighting rhythm intensity as a universally protective factor alongside more individualized vulnerabilities in timing and regularity, offers a foundation for precision prevention. At the population level, interventions that enhance rhythm intensity, such as structured physical activity programs, may broadly reduce aging risk. In contrast, individualized strategies that address phase misalignment or rhythm regularity, circadian-aligned nutrition and behavioral scheduling, hold promise for mitigating personal vulnerabilities. Importantly, because these digital phenotypes can be passively derived from consumer-grade wearable devices, scalable, ecologically valid, and continuous monitoring of circadian health is now achievable, paving the way toward personalized circadian medicine.

Several limitations warrant consideration. First, the AoURP cohort with Fitbit data was predominantly young, female, white, and college-educated^53^. In addition, participants provided their own devices, and the middle-aged to older adults in this cohort were found more physically active than the national average^53^. Despite this relatively healthy and high-activity profile, we still observed robust associations between rest-activity rhythm and biological aging, suggesting that even stronger associations may be present in more sedentary or clinically vulnerable populations. Validation study in more heterogeneous cohorts across diverse demographic, socioeconomic, and environmental contexts is therefore needed. Second, reverse causality cannot be excluded given the observational design, although we attempted to mitigate this concern by testing time-varying effects. Third, Fitbit step counts may underestimate non-stepping activities such as cycling or resistance training^53^. In addition, day-level valid-wear criterion based on total steps and device-reported wear time does not capture intra-day nonwear bouts. Future work is warranted to incorporate raw accelerometry to derive more precise activity characterization and develop methodologies for consumer device-specific nonwear algorithm, improving exposure ascertainment and rest-activity rhythm estimates. Finally, we acknowledge the limitations of EHR-based outcome ascertainment. Certain events, such as laboratory assessments performed outside routine clinical encounters, may not be fully captured. Nevertheless, prior studies have demonstrated the validity of EHR data for estimating biological age measures^18,54^, supporting the reliability of our approach.

Despite these limitations, our study brings several notable strengths. Unlike prior investigations that relied on research-grade accelerometers, cross-sectional measurements, or short-term follow-up, our analysis leveraged commercially available devices, a large-scale prospective cohort, and multi-year monitoring periods, thereby enhancing translational potential to real-world applications. In addition, a substantial portion of Fitbit data was collected before participants formally enrolled in AoURP, minimizing observer effects^53^ and reflecting free-living rest-activity patterns. Methodologically, by jointly quantifying between-person variability and within-person temporal stability, we demonstrated that 24-hour rhythm estimates captured both population-level associations and individual-level longitudinal robustness. Moreover, sex-stratified models allowed us to rigorously examine sex differences in circadian-aging association, revealing nuanced, biologically plausible activity patterns. Finally, strict data quality control, extended longitudinal monitoring, and the integration of both scalar metrics and functional principal components provide a comprehensive framework for deep digital phenotyping.

In summary, this study demonstrates that circadian rhythmicity exerts a lasting influence on biological aging trajectories. By leveraging both conventional CRAR metrics and functional data analysis, we show that rhythm intensity offers stable, universal protection, while timing and fragmentation introduce sex-specific vulnerabilities. These insights elevate circadian rhythmicity from a descriptive phenomenon to a digital biomarker of aging, opening a pathway toward scalable, personalized, and temporally targeted interventions. If future trials confirm that modifying these digital phenotypes can alter aging trajectories, wearable-guided circadian interventions may redefine preventive medicine for healthy longevity.

## METHODS

### Study participants

We focused on participants from the *All of Us* Research Program (AoURP), a prospective, longitudinal cohort study in the United States funded by the National Institutes of Health, which aims to enroll at least one million individuals and create a rich research biobank integrating diverse health data, including electronic health records (EHR), biospecimens, surveys, wearables, and medical imaging^41–43^. As of 2024, AoURP had enrolled more than 800,000 participants, providing a uniquely comprehensive resource for multi-modal, population-level longitudinal research. For this study, we used registered and controlled tier data available on the AoU Researcher Workbench v8.

From this cohort, we identified 34,217 participants who had contributed Fitbit data through the Bring Your Own Device (BYOD) program, consented to share their EHR data, and provided valid physical measurements (**Supplementary** Fig. 6). We excluded individuals younger than 18 years during the monitoring period or did not meet valid wear day criteria (refer to Fitbit data acquisition and processing section). To ensure continuous and stable assessment of circadian rest-activity rhythmicity, only participants with complete step data for a full 12-month period within a given calendar year were retained. Among the 11,698 participants meeting these criteria, we estimated phenotypic age for each calendar year using linked EHR data. The final analytic cohort consisted of 2,222 participants with multi-year valid Fitbit data and corresponding biological age estimates, contributing 8,447 person-years of follow-up.

### Fitbit data acquisition and processing

Participants linked their personal Fitbit accounts to the AoURP Participant Portal through the BYOD program, authorizing access to both prospective and historical data. All data were de-identified according to AoURP privacy protocols, with direct identifiers removed and all date-time fields randomly shifted by 1 to 365 days. Step count data were obtained at both daily and intraday resolutions, with intraday time series used to quantify daily activity patterns and compute rest-activity rhythm metrics. Valid wear days were defined as at least 10 hours of wear time and a minimum of 100 steps, consistent with prior validation studies^53,58^. Fitbit devices have been shown to provide accurate estimates of physical activity and circadian rhythm metrics^59^ compared with research-grade accelerometers.

### Biological age estimation

The primary outcome was phenotypic age (PhenoAge), a validated biomarker-based estimate of biological age that predicts morbidity, functional decline, and all-cause mortality^30^. PhenoAge has also been associated with wearable-derived rhythmicity in prior cross-sectional analyses^16–18^. It is calculated using the Gompertz proportional hazards model trained on nine clinical biomarkers, including albumin, alkaline phosphatase, creatinine, C-reactive protein (CRP), glucose, mean cell volume, red cell distribution width, white blood cell count, and lymphocyte percentage, along with chronological age^30,60^.

Annual biomarker values were extracted from linked EHR records for each year of Fitbit monitoring (**Supplementary Table 4**). Biomarker units were harmonized to match the original PhenoAge algorithm. Outliers exceeding five standard deviations (SD) from the mean were manually reviewed and excluded. When up to two biomarkers were missing, values were imputed using the cohort mean; for CRP, which exhibited >50% missing values, the global mean was substituted. This imputation has been shown to exert minimal influence on downstream analyses.

To examine associations between rest-activity rhythms and biological aging, we calculated biological age acceleration (ΔAge) as the residual of PhenoAge regressed on chronological age^61^. Participants with ΔAge>0 were classified as fast aging, and those with ΔAge≤0 as slow aging. The final analytic cohort included only individuals with at least two valid PhenoAge measurements temporally aligned with years of valid Fitbit monitoring, thus allowing to examine longitudinal relationships.

### Assessment of circadian rest-activity rhythms and functional principal components

Circadian rest-activity rhythm (CRAR) metrics were derived using parametric and non-parametric approaches^50–52^ and data-driven functional principal component analysis^64^. For parametric estimation, we applied cosinor analysis to step count time series to quantify the intensity and timing of circadian rhythmic fluctuations. The cosinor model has been applied to analyze circadian patterns from melatonin, core body temperature, and wearable-derived rest-activity rhythm^54–56^. Specifically, a 24-hour single-component cosinor model was fitted to each individual’s data to estimate the MESOR (Midline Estimating Statistic of Rhythm), amplitude (half the range of rhythmic variation within the cycle), and acrophase (timing of peak activity)^68,69^. Model fitting used an iterative least-squares procedure, assuming a fixed circadian period of 24 hours, consistent with prior work^18^.

We derived seven non-parametric CRAR metrics that have been significantly associated with aging-related outcomes^17,59–61^. These metrics include M10, M10 start time, L5, L5 start time, relative amplitude (RA), interdaily stability (IS), and intradaily variability (IV). M10 represents the mean activity during the most active 10 consecutive hours within a 24-hour period and serves as a measure of daytime activity intensity. L5 denotes the average activity during the least active 5 consecutive hours, typically corresponding to the nighttime rest period. The timing of these periods is captured by M10 start time and L5 start time, defined as the clock time at which the M10 and L5 windows begin, respectively. These timing variables provide non-parametric estimates of activity phase. RA, calculated as (M10−L5)/(M10+L5), measures rhythm strength, with higher values indicating greater separation between active and rest phases and lower values suggesting dampened or flattened rhythmicity.

We quantified the degree of rhythm regularity using IS and IV metrics. IS reflects the degree of consistency in activity patterns over multiple days^63,72^, comparing the variance of mean activity at each clock time across days to the total variance. A higher IS value indicates greater day-to-day regularity, whereas a lower IS suggests irregular, unstable daily patterns. IS ranges from 0 to 1. IV captures the degree of fragmentation in activity within a 24-hour period. It is calculated as the ratio of the variance of successive differences in activity to the overall variance in daily activity. Lower IV values reflect more consolidated rhythms with distinct separation between active and rest periods, while higher IV values indicate frequent transitions between activity and inactivity, suggestive fragmented rhythm. IV ranges from 0 to 2. All CRAR metrics were computed from 10-minute epoch data, aggregated monthly, and then averaged across each calendar year.

To characterize higher-order latent patterns in daily activity profiles beyond conventional circadian rhythm metrics, we applied functional principal component analysis (fPCA)^64^. Step count trajectories were treated as functional data as daily activity follows a continuous and smooth temporal process^73,74^. For each participant-year, step counts were aggregated into 5-minute epochs, yielding 288 observations per day. We then projected these trajectories onto a nine-term Fourier basis, consistent with prior studies that effectively captures diurnal periodicity with minimal overfitting^74,75^. Empirical evaluation of 7-10 harmonics demonstrated that harmonics beyond nine terms provided only marginal gains in variance explained while increasing model complexity. The resulting smoothed functions were subjected to fPCA, which decomposes the covariance matrix into an orthonormal basis of eigenfunctions that capture principal components modes of variation. We retained the first four functional principal components (fPCs), which jointly explained 88% of the total variance. Each participant-year was represented by a vector of four orthogonal fPC scores. These scores represent distinct and interpretable dimensions of behavioral rhythmicity at the population level. For consistency across models, each fPC score was standardized and categorized into quartiles for regression and mixed-effects analyses.

### Covariates

We pre-specified a comprehensive set of covariates based on prior evidence and theoretical considerations to account for potential confounding in statistical models^16,18,53,76^. At initial enrollment, demographic and socioeconomic data were collected using self-identified information provided by participants^77^. Demographic variables included chronological age at the time of Fitbit monitoring (in years), sex assigned at birth (male or female), and race/ethnicity (White, Black, or Other). Socioeconomic status was assessed through educational attainment (categorized as college or above, high school graduate or GED, and missing/other), employment status (employed, retired, unemployed, or other), and annual household income (US dollars), categorized as high (annual income >$100,000), medium (between $50,000 and $100,000), low (<$50,000), or missing. Body-mass index (BMI) was calculated from measurements of height (m) and weight (kg) using weight divided by height^2^ in physical measurements data obtained closest in time to the Fitbit data. BMI categories were divided by as normal or underweight (<25 kg/m²), overweight (25-29.9 kg/m²), or obese (≥30 kg/m²). Sleep duration was obtained from Fitbit sleep data by averaging the main sleep episode per year during the monitoring period. Smoking history was self-reported using concept code 1585857 (“yes” if lifetime cigarette use exceeded 100 cigarettes, otherwise “no”), and alcohol consumption was coded as a binary variable using concept code 1586198. The presence of major comorbidities, including cardiovascular disease (CVD), type 2 diabetes, and cancer, was ascertained from structured EHR data using predefined International Classification of Diseases (ICD) codes (**Supplementary Table 5**). In the linear mixed-effects models, we accounted for time-varying covariates where appropriate.

Specifically, age, BMI, and comorbidity status (presence or absence of CVD, type 2 diabetes, and cancer) were treated as time-varying and matched to each participant-year based on the corresponding year of Fitbit and biological age data.

### Statistical analysis

Baseline characteristics were summarized as means and SDs for continuous variables and counts with percentages for categorical variables. To examine cross-sectional associations between CRAR metrics or fPC scores and accelerated biological aging at both baseline and 5-year follow-up, we fitted generalized linear models with a logit link, adjusting for all pre-specified covariates. To evaluate longitudinal associations between activity metrics and biological age acceleration trajectories, we employed linear mixed-effects models with ΔAge as the continuous, time-dependent outcome, the metric of interest as a fixed effect, and participant-level random intercepts to account for individual heterogeneity and correlation induced by repeated measures. Models were adjusted for age, sex, race/ethnicity, BMI, and comorbidity status as time-varying covariates.

To evaluate the robustness of our findings, we performed multiple sensitivity analyses. First, sex-stratified analyses were performed, repeating the regression models separately for male and female (**Supplementary Tables 1-2**) and by conducting sex-stratified fPCA (**Supplementary** Fig. 4). Second, we tested for time-by-metric interaction effects in the linear mixed effect models to assess whether the associations between behavioral metrics and biological aging varied over follow-up period (**Supplementary Table 3**).

To enable direct comparison of effect sizes across heterogeneous metrics, all CRAR and fPC variables were categorized into quartile groups prior to model fitting. This facilitated consistent quartile definitions across metrics, while model coefficients represented differences in outcome between quartile categories relative to the reference group.

Consistent with the AoURP Data and Statistics Dissemination Policy, model results with cell sizes ≤20 observations were not reported. All analyses were conducted in Python (version 3.9) within the AoURP Workbench, using parallel processing on four nodes when applicable. Statistical significance was defined as a two-sided p-value <0.05.

## DATA AVAILABILITY

Data supporting the results in this study are available for approved researchers following registration, completion of ethics training, and attestation of a data use agreement through the *All of Us* Research Workbench platform.

## CODE AVAILABILITY

The codes that support the findings of this study are available for approved researchers on the *All of Us* Research Workbench platform by contacting to the corresponding author.

## Data Availability

Data supporting the results in this study are available for approved researchers following registration, completion of ethics training, and attestation of a data use agreement through the All of Us Research Workbench platform.

## ACKNOWLEDGEMENTS

This study received no funding.

## AUTHOR CONTRIBUTIONS

J.S. contributed conceptualization, methodology, data curation, formal analysis, writing (original draft), and writing (review and editing). J.P.O. contributed methodology, data interpretation, and writing (review and editing).

